# RT-PCR/MALDI-TOF diagnostic target performance reflects circulating SARS-CoV-2 variant diversity in New York City

**DOI:** 10.1101/2021.12.04.21267265

**Authors:** Matthew M. Hernandez, Radhika Banu, Ana S. Gonzalez-Reiche, Brandon Gray, Paras Shrestha, Liyong Cao, Feng Chen, Huanzhi Shi, Ayman Hanna, Juan David Ramírez, Adriana van de Guchte, Robert Sebra, Mount Sinai PSP Study Group, Melissa R. Gitman, Michael D. Nowak, Carlos Cordon-Cardo, Ted E. Schutzbank, Viviana Simon, Harm van Bakel, Emilia Mia Sordillo, Alberto E. Paniz-Mondolfi

**Affiliations:** Department of Pathology, Molecular, and Cell-Based Medicine, Icahn School of Medicine at Mount Sinai, New York, NY 10029, USA; Department of Genetics and Genomic Sciences, Icahn School of Medicine at Mount Sinai, New York, NY 10029, USA; Centro de Investigaciones en Microbiología y Biotecnología-UR (CIMBIUR), Facultad de Ciencias Naturales, Universidad del Rosario, Bogotá, Colombia; Icahn Institute for Data Science and Genomic Technology, Icahn School of Medicine at Mount Sinai, New York, NY 10029, USA; Black Family Stem Cell Institute, Icahn School of Medicine at Mount Sinai, New York, NY 10029, USA; Sema4, a Mount Sinai venture, Stamford, CT 06902, USA; Department of Microbiology, Icahn School of Medicine at Mount Sinai, New York, NY 10029, USA; Senior Scientific Affairs Manager, Infectious Diseases, Agena Bioscience, San Diego, CA 92121, USA; Division of Infectious Diseases, Department of Medicine, Icahn School of Medicine at Mount Sinai, New York, NY 10029, USA; The Global Health and Emerging Pathogens Institute, Icahn School of Medicine at Mount Sinai, New York, NY 10029, USA

**Keywords:** RT-PCR, MALDI-TOF, SARS-CoV-2, Delta, variants, diagnostic, dropout

## Abstract

As severe acute respiratory syndrome coronavirus 2 (SARS-CoV-2) continues to circulate, multiple variants of concern (VOC) have emerged. New variants pose challenges for diagnostic platforms since sequence diversity can alter primer/probe binding sites (PBS), causing false-negative results. The Agena MassARRAY^®^ SARS-CoV-2 Panel utilizes reverse-transcription polymerase chain reaction and mass-spectrometry to detect five multiplex targets across *N* and *ORF1ab* genes. Herein, we utilize a dataset of 256 SARS-CoV-2-positive specimens collected between April 11, 2021-August 28, 2021 to evaluate target performance with paired sequencing data. During this timeframe, two targets in the *N* gene (N2, N3) were subject to the greatest sequence diversity. In specimens with N3 dropout, 69% harbored the Alpha-specific A28095U polymorphism that introduces a 3’-mismatch to the N3 forward PBS and increases risk of target dropout relative to specimens with 28095A (relative risk (RR): 20.02; p<0.0001; 95% Confidence Interval (CI): 11.36-35.72). Furthermore, among specimens with N2 dropout, 90% harbored the Delta-specific G28916U polymorphism that creates a 3’-mismatch to the N2 probe PBS and increases target dropout risk (RR: 11.92; p<0.0001; 95% CI: 8.17-14.06). These findings highlight the robust capability of Agena MassARRAY^®^ SARS-CoV-2 Panel target results to reveal circulating virus diversity and underscore the power of multi-target design to capture VOC.

## Introduction

Since the recognition and global spread of coronavirus disease 2019 (COVID-19), nucleic acid amplification testing has been the diagnostic mainstay for identifying new severe acute respiratory syndrome coranvirus-2 (SARS-CoV-2) infections. Historically, the majority of diagnostic target primers and probes have been designed from viral genomes characterized early in the pandemic. However, in the two years since the first SARS-CoV-2 genome was identified, viral variants have emerged in response to various evolutionary pressures ^1,2^. Emergence of variants of concern (VOC) and of interest (VOI) poses a hurdle for diagnostic assays as sequence variation generates mismatches to primer/probe binding sites (PBSs), potentially causing target dropout and false-negative results ^3,4^. Failure to promptly detect infections with emerging variants that may be associated with greater transmissibility and infectivity represents a substantive risk for forward transmission in the community.

The diagnostic assays that are most robust for detection of SARS-CoV-2 and least affected by viral sequence changes are designed to amplify two or more targets across different viral genes (e.g., open reading frame 1ab (*ORF1ab*), nucleocapsid (*N*), spike (*S*), and envelope (*E*)). This redundancy can compensate for individual target dropout due to inherent biochemical differences and analytical sensitivities among targets or due to emergent viral genome diversity at PBS sequences. Several groups have demonstrated the utility of platforms with multiple targets to accurately detect emerging SARS-CoV-2 variants ^3,5,6^. Furthermore, target performance patterns on these platforms can be used to elucidate dynamics of circulating variants in communities. Indeed, multiple reports described the use of dropout of diagnostic targets designed at *S* ^5,7,8^ or *N* ^9^ genes as a screen for PBS-specific polymorphisms that have arisen among emergent SARS-CoV-2 viruses such as the Alpha variant (B.1.1.7 lineage).

In early 2021, the Delta variant (B.1.617.2 lineage) of SARS-CoV-2 surged in India^10–12^, and soon Delta and its sublineages (AY.x) quickly spread across the globe ^2,13–15^. By Fall 2021, despite increased vaccination rates and infection prevention methods ^16–18^, Delta and its sublineages became predominant in New York City (NYC) and abroad ^10,13,14^. Given that emergent variants may be associated with resistance to therapeutic antibodies ^19,20^, increased transmissibility ^12,16,21,22^, or reduced vaccine effectiveness ^16,17,23,24^, diagnosing new infections is vital. Several mutation-specific assays have been designed to detect lineage-specific *S* gene mutations in Delta and older variants ^25–27^. However, few assays have been evaluated for their ability to capture contemporary variants in routine clinical settings, or to shed light on sequence diversity in virus genes other than Spike (S).

We recently reported the power of the Agena MassARRAY® SARS-CoV-2 Panel for detection of emergent SARS-CoV-2 variants in NYC during the first months of 2021 and the utility of diagnostic target performance to illuminate sequence diversity among then-predominant Alpha variant genomes ^28^. This assay combines reverse-transcription polymerase chain reaction and matrix-assisted laser desorption-ionization time-of-flight mass spectrometry (RT-PCR/MALDI-TOF) to detect five targets in the SARS-CoV-2 genome. Coincident with the spread of the Delta VOC in NYC, we observed a distinct change in the target performance patterns that reflect ongoing viral evolution, prompting us to examine the corresponding genomic sequence data.

For the current study, we compared paired RT-PCR/MALDI-TOF diagnostic target data and genome sequencing data for SARS-CoV-2-positive specimens that were collected from April 11, 2021 – August 28, 2021, and recovered, deidentified, and sequenced as part of the Mount Sinai Health System (MSHS) Pathogen Surveillance Program (PSP) at the Icahn School of Medicine at Mount Sinai (ISMMS). Herein, we utilize a dataset of 256 genomes from SARS-CoV-2-positive clinical specimens to identify PBS mismatches in the *N* gene and lineage-specific sequence diversity that impacted target performance patterns on the Agena MassARRAY^®^ SARS-CoV-2 Panel.

## Materials and Methods

### Ethics statement

This study was reviewed and approved by the Institutional Review Board of the ISMMS (HS#13-00981).

### SARS-CoV-2 specimen collection and testing

Upper respiratory tract (e.g., nasopharyngeal, anterior nares) and saliva specimens collected between April 11, 2021 – August 28, 2021 for SARS-CoV-2 diagnostic testing in the Molecular Microbiology Laboratories (MML) of the MSHS Clinical Laboratory, which is certified under Clinical Laboratory Improvement Amendments of 1988, 42 U.S.C. §263a and meets requirements to perform high-complexity tests were eligible for inclusion in this study. Overall, 177,059 upper respiratory and saliva specimens underwent SARS-CoV-2 clinical testing at MSHS MML during the study period, and subset of 256 were deidentified and underwent SARS-CoV-2 next-generation sequencing as previously described ^28–30^. For this retrospective study, we utilized deidentified data for diagnostic specimens tested on the Agena MassARRAY^®^ SARS-CoV-2 Panel and MassARRAY^®^ System (Agena, CPM384) platform. Details of processing and SARS-CoV-2 testing of upper respiratory and saliva specimens have been described previously ^28,31^.

This RT-PCR/MALDI-TOF assay detects five viral targets: three in the nucleocapsid (*N*) gene (N1, N2, N3) and two in the *ORF1ab* gene (ORF1A, ORF1AB). Results were interpreted as positive if ≥ 2 targets were detected or negative if < 2 targets were detected. If no MS2 internal extraction control (IC) and no targets were detected, the result was invalid and required repeat testing of the specimen before reporting. If IC was detected and no targets were detected, the sample was interpreted as negative.

### SARS-CoV-2 sequencing, assembly and phylogenetic analyses

SARS-CoV-2 viral RNA underwent reverse-transcription, PCR amplification and next-generation sequencing followed by genome assembly and lineage assignment using a phylogenetic-based nomenclature as described by Rambaut et al. ^32^ using the Pangolin v3.1.14 tool and PANGO-v1.2.81 nomenclature scheme ^33^ as previously described ^29,30^. Ultimately, this yielded 244 complete genomes (≥95% completeness) and 12 partial genomes (<95% completeness).

### Agena target sequence alignment

Agena MassARRAY^®^ target detection results were matched to the corresponding genome sequences. Primer and probe sequences for each Agena target were obtained from published FDA EUA documentation for the Agena MassARRAY^®^ SARS-CoV-2 Panel (**Table S1**) ^34^. We generated reverse-complement sequences for reverse primers for all five targets and probes that are designed in the reverse orientation (e.g., N1-N3). An unaligned FASTA file including sequence data for the clinical specimens and the Wuhan-Hu-1 reference sequence (NCBI nucleotide: NC_045512.2 (Genbank: MN908947.3)) was generated for each of the fifteen primers/probes. Alignment of specimen sequences with each primer/probe sequence was performed using the Multiple Alignment using Fast Fourier Transform (MAFFT) platform ^35,36^ as previously described ^28^. To enable inclusion of incomplete genomes that had intact regions sequenced at PBSs, we did not remove uninformative sequences (e.g., with ambiguous letters). Otherwise, the default settings were used to align all sequences to the reference genome, which generated a resulting FASTA alignment file for each primer and probe sequence.

### Sequence variation in primer/probe target regions

To examine sequence diversity at each primer/probe PBS region, nucleotide diversity per basepair was calculated using the PopGenome population genomics analysis package in R (v4.1.2) ^37^. Nucleotide diversity per site (π) was calculated by dividing the nucleotide diversity of all sequences within each PBS sliding window by the number of nucleotide sites in each corresponding PBS.

To identify and calculate mismatch frequency at each PBS, custom Unix-code ^38^ was used to identify mismatches at each nucleotide position within each primer and probe sequence as descried previously ^28^. PBS mismatch frequency by position of forward/reverse primer and probe sequences were calculated on Microsoft Excel v16.54. To account for differences in completeness of consensus genomes, the number of PBS mismatches was normalized to the number of nucleotides in the PBS of each specimen consensus sequence.

To compare global sequence data for lineages and polymorphisms relevant to this study, we queried publicly-available sequence data from the Global Initiative on Sharing Avian Influenza Data (GISAID) EpiCoV database through their online user interface (https://www.epicov.org, accessed October 16, 2021). Using available filters for collection date ranges, amino acid substitutions, and variant classification, we calculated the frequencies of distinct genomes deposited. Specifically, to distinguish nucleotide polymorphisms at SARS-CoV-2 position 28916, we identified the number of variations at the corresponding 215^th^ amino acid in the N protein. We determined nucleotide changes by comparing the codon of the native glycine amino acid (28916-GGU-28918) to the codons of the variant amino acids. These changes (nucleotide change underlined) included cysteine (<u>U</u>GU), serine (<u>A</u>GU), and arginine (<u>C</u>GU) amino acids.

### Statistical analyses

For statistical comparison of fraction of PBS with mismatches in genomes with detected targets versus those with dropout, normality was assessed by D’Agostino and Pearson test (GraphPad Prism v9.2.0). All distributions were non-parametric, and thus, a Mann-Whitney test (two-tailed) was performed (GraphPad). To determine if specific mismatches were associated with specific target dropout results, specimens were grouped by (1) presence or absence of the mismatch of interest (in the setting of no other mismatches) and (2) detection or dropout of the target of interest – which resulted in a 2×2 contingency table which underwent association testing by Fisher’s exact test. In addition to p-value, relative risk (RR) ratios and 95% confidence intervals (CI) are provided for association testing.

### Display Items

All figures are original and were generated using the GraphPad Prism software 9.2.0, Microsoft Excel v16.54, and finished in Adobe Illustrator 2021 (v.25.4.1). Alignment plots were developed using the package ggmsa (v.1.1.0) on R ^39^ and finished in Adobe Illustrator.

## Results

Of 448 SARS-CoV-2-positive specimens, 117 (26%) had all five targets detected with the remaining having one (n=247) or more (n=84) targets dropout. For the subset of 256 SARS-CoV-2-positive specimens sequenced in our study, all five diagnostic targets were detected in 96 (38%), with the remaining having one (n=140) or more (n=20) targets dropout (**Table S2**).

When we calculated weekly detection rates for individual targets among these SARS-CoV-2-positive specimens, the N2 (0.61) and N3 (0.77) targets had the lowest average detection rates (**Figure 1A**). Notably, N2 and N3 detection rates were lowest at distinct periods of time. Among weeks with ≥3 SARS-CoV-2-positive specimens sequenced, the N2 detection rate was lowest (0.13) in the week ending July 31, and the N3 detection rate was lowest (0.41) in the week ending May 1. We hypothesized this shift was due to changes in the population of circulating SARS-CoV-2 variants. When we compared individual target detection rates to reported prevalence of variants in NYC during the same time period, we found the N3 detection rate was lowest when the Alpha variant (B.1.1.7 + Q.x) was predominant at 33% (**Figure 1B**). Similarly, the N2 detection rate was lowest when the Delta variant (B.1.617.2 + AY.x) comprised 96% of circulating SARS-CoV-2 in NYC. Given these findings, we used diagnostic target and paired genome sequence data to identify mismatches to Agena MassARRAY^®^ target PBSs to determine impact of SARS-CoV-2 genomic diversity on diagnostic target performance.

**Figure 1.**
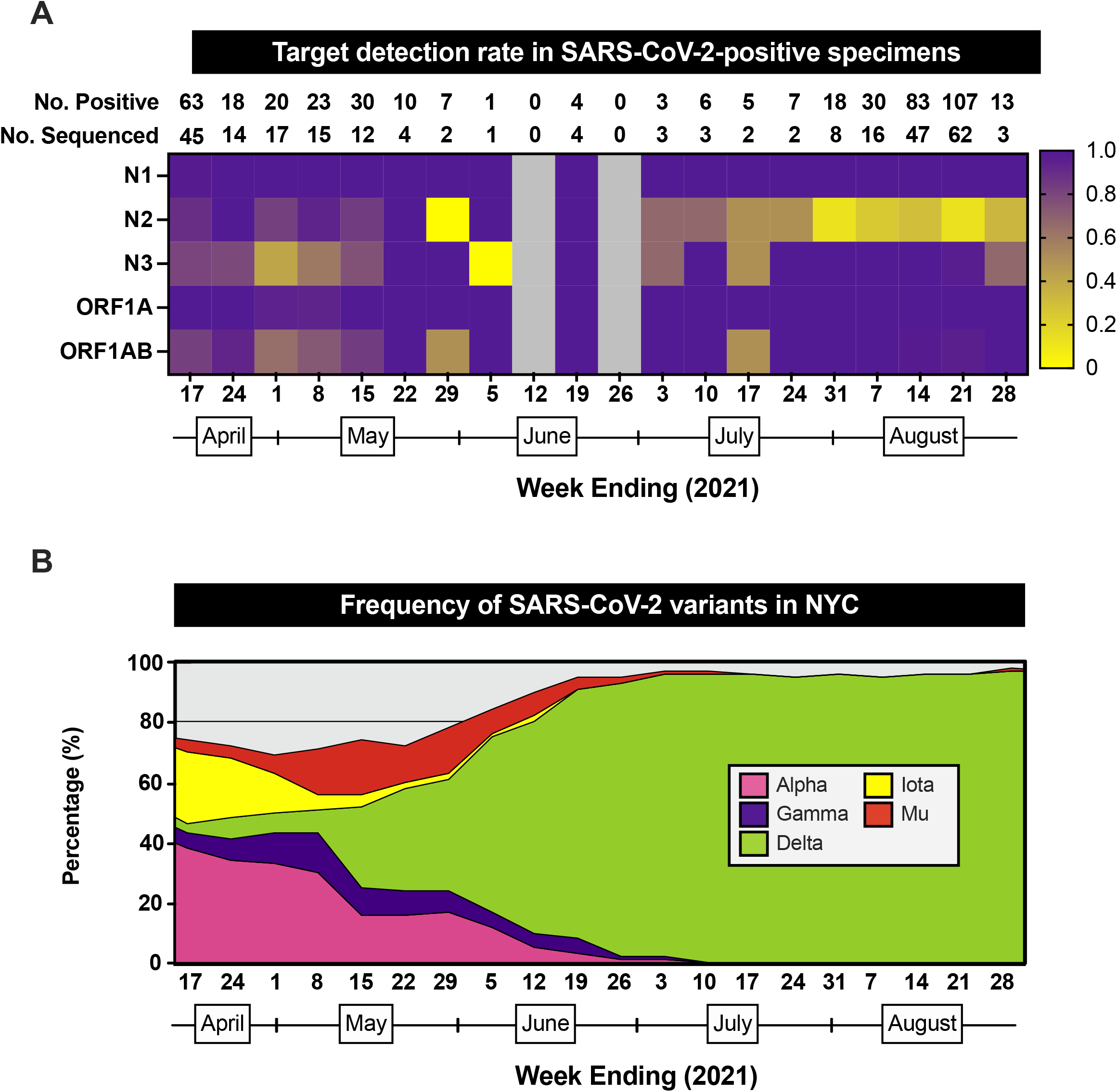
RT-PCR/MALDI-TOF target detection rate in SARS-CoV-2-positive specimens. (A) Heatmap depicting the proportion of SARS-CoV-2-positive specimens that have detectable RT-PCR/MALDI-TOF targets (N1, N2, N3, ORF1A, ORF1AB) by week from April 11 through August 28, 2021. The total number of SARS-CoV-2-positive specimens and the number of SARS-CoV-2-positive specimens sequenced by pathogen surveillance are depicted above each week (column). Grey boxes indicate weeks where no specimens were positive for SARS-CoV-2 on this platform. (B) Stacked area plots depicting frequencies of SARS-CoV-2 variants reported by publicly-available NYC Department of Health surveillance data within the same timeframe.

### Impact of mismatches on target performance

We aligned each diagnostic target PBS to each of the 256 SARS-CoV-2 genome sequences and calculated the number of mismatches to each target PBS for both detected and undetected targets (**Figure 2**). For four targets (N1, N3, ORF1A, ORF1AB), when the individual target was detected there was perfect complementarity (0 mismatches) between genome sequences and forward, reverse, and probe sequences for almost all (>99%) specimens. However, for specimens in which the N2 target was detected, mismatches occurred both at N2 forward (34.3%) and reverse (46.6%) PBSs (**Figure 2B**). Specifically, among specimens with detectable N2 target, 1-3 mismatches to the forward PBS and up to 1 mismatch to the reverse PBS were observed.

**Figure 2.**
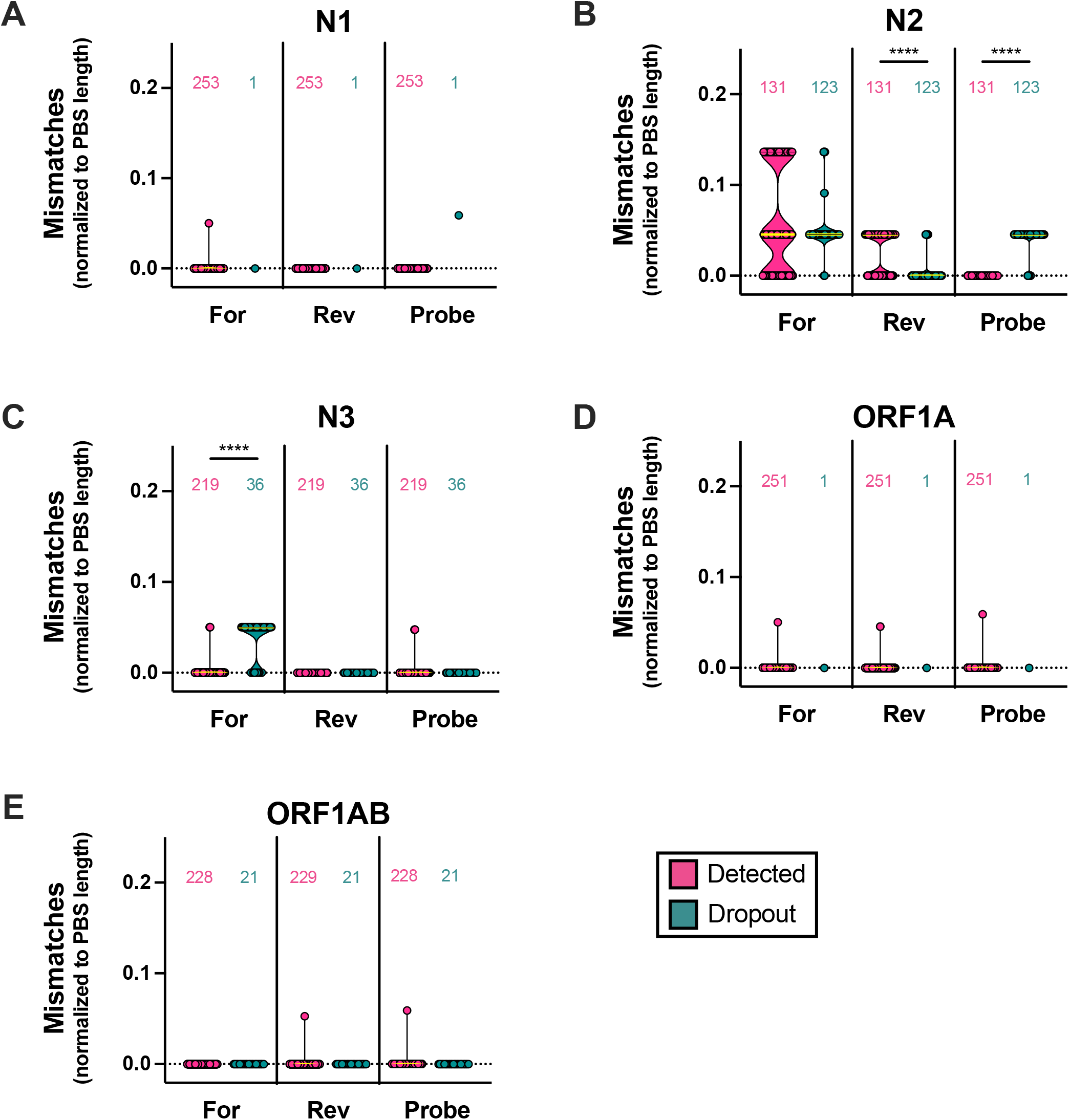
Frequencies of target PBS mismatches and RT-PCR/MALDI-TOF target detection results. Number of mismatches normalized to the number of nucleotides in primer/probe binding sites (PBS length) across five diagnostic targets: (A) N1, (B) N2, (C) N3, (D) ORF1A, (E) ORF1AB. Each point represents the calculated mismatches per specimen consensus genome for each target PBS. Violin plots represent the distribution as density of the points grouped by primer/probe sequence (forward (For), reverse (Rev), Probe) and by target detection result (detected (magenta), dropout (turquoise)). Numbers of genomes analyzed for mismatches are depicted above each violin plot. Medians (yellow lines) are depicted and bars above distributions represent statistical comparisons (Mann-Whitney test; *, P<0.05; ****, P<0.0001).

Notably, specimens with N2 target dropout had a significantly greater number of mismatches to the N2 probe PBS (**Figure 2B**). The same was found for specimens with N3 dropout and mismatches to the N3 forward PBS (**Figure 2C**). Of interest, more mismatches to the N2 reverse PBS were found for specimens in which the N2 target was detected than for those with N2 dropout. Indeed, the majority of specimens with detectable N2 target (85%) harbored mismatches in N2 forward or reverse PBSs, but 0% of specimens with detectable N2 target harbored mismatches to the N2 probe PBS.

### Site-specific polymorphisms affect target performance

Because mismatch position in PBSs can affect primer/probes binding to target sites ^40–43^, we characterized mismatch frequencies by position in each PBS (**Figure 2**). At least one mismatch was identified in every target PBS with the exception of the N1 reverse, N3 reverse, and the ORF1AB forward primers which had zero mismatches to all possible haplotypes in our dataset. To assess the level of sequence diversity across each target, we calculated the nucleotide diversity per site (π) for each PBS sequence. Overall, genomes sequenced in this timeframe demonstrated the most diversity at N2 target PBSs (≥0.01260) followed by the N3 forward PBS (0.01075) (**Figure 3B-C**). Diversity at these targets was at least seven-fold more than that at all other target PBSs. Notably, this diversity corresponded with a predominance of Delta (56.7%), Alpha (15.7%),, and Iota (16.5%) variant genomes in this dataset.

**Figure 3.**
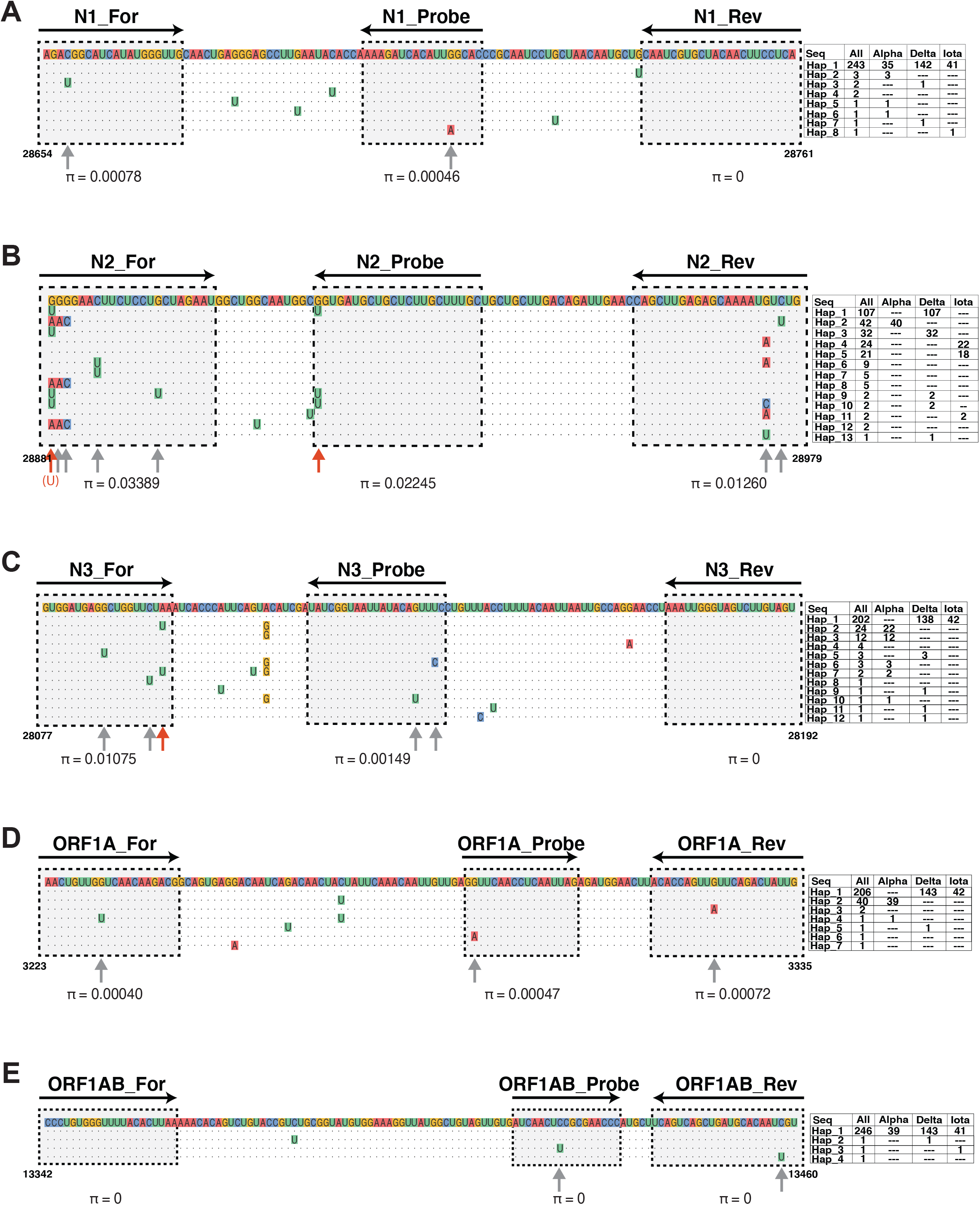
Alignment of SARS-CoV-2 haplotypes to RT-PCR/MALDI-TOF targets. Multiple sequence alignment of haplotype sequences to forward, reverse, and probe binding sites for (A) N1, (B) N2, (C) N3, (D) ORF1A, and (E) ORF1AB targets. In each alignment, the reference sequence (NC_045512.2) is annotated above comprised of color-coded nucleotides. The coordinates of the reference sequence are annotated at the bottom corners of each alignment. Each row represents a haplotype sequence that aligns to the target site. Absolute counts of each haplotype (e.g., Hap_1, Hap_2, etc.) within the dataset and counts stratified by variant lineage (e.g., Alpha, Delta, Iota) are depicted to the right side of each haplotype sequence. Positions of primer/probe sequences are indicated by arrows where tail to arrowhead reflects 5’ to 3’ directionality. Grey boxes with black dotted borders outline PBSs. Dots represent conserved nucleotides at each position and mismatched nucleotides are indicated. PBS mismatches are indicated by arrows; red arrows reflect mismatches that are significantly associated with target dropout. Notably, the association of the G28881U mismatch to the N2 forward PBS (panel C, red “(U)”) with target dropout is distinguished from the G28881A mismatch. Nucleotide diversity (π) of sequences at each PBS is indicated.

To identify individual mismatches associated with target dropout, we measured the proportion of genomes with a mismatch at each PBS position and compared this level across specimens that yielded target detection or dropout (**Figure S1**). From 5’-to-3’ direction, there were mismatches to 1^st^ – 3^rd^ bp of the N2 forward primer PBS (SARS-CoV-2 genome positions 28881 – 28883) in genomes that yielded both N2 target detection and dropout (**Figure 3B, Figure S1B**). Overall, there were 192 specimens with at least one mismatch to the first 3 bp of the N2 forward primer. Of these, 49 (26%) had a concurrent trio of substitutions in the *N* gene: G28881A, G28882A, and G28883C; the remaining 143 (74%) harbored a single G28881U substitution. The GGG-to-AAC and the G28881U substitutions were independent of one another and no combinations of the two were detected. The majority of specimens with the trio (88%) had detectable N2 target. In contrast, 78% of specimens with G28881U resulted in dropout which – despite its position at the 5’-end of the N2 forward primer – represents a significant association of G28881U with N2 target dropout (p<0.0001).

Interestingly, of 123 genomes with N2 target dropout we identified 111 (90%) that harbored a mismatch at the terminal 3’ base of the 22-bp-long N2 probe (**Figure 3B, Figure S1B**). All mismatches are the result of a guanine-to-uracil substitution in the *N* gene (G28916U). This substitution was not identified among any of the 131 specimens with detectable N2 target. In fact, the relative risk of N2 target dropout in the setting of the G28916U substitution was 11.9-times greater than genomes with the native guanine (p<0.0001; 95% CI: 8.17-14.06). We also determined that all 111 genomes with the G28916U substitution have the G28881U substitution. The linkage of the G28881U and G28916U polymorphisms likely explains why the former – a 5’-end mismatch to the N2 forward PBS – is associated with N2 target dropout. Indeed, 100% of genomes (n=32) that possess the G28881U substitution alone resulted in N2 target detection.

In the current study, of 36 specimens with N3 target dropout, 25 (69%) had a mismatch at the penultimate position of the 3’-end of the 20-bp-long N3 forward primer (**Figure 3C, Figure S1C**). This mismatch corresponded with the adenine-to-uracil substitution in the *ORF8* gene (A28095U) which causes a 20-fold risk of N3 target dropout relative to genomes with the native nucleotide (p<0.0001; 95% CI: 11.36-35.72) which is consistent with our findings earlier in the pandemic ^28^, prior to the current study period.

We assessed whether the associations between mismatch and corresponding target dropout persist when quantity of virus genomic material in specimens is controlled as previously described ^28^. When analyses are limited to specimens for which all non-N2 targets are detected, the association of the G28916U substitution with N2 dropout remains significant (p<0.0001; RR: 20.2; 95% CI: 14.58-47.48). In addition, when analyses are limited to specimens for which all non-N3 targets are detected, the A28095U substitution remains significantly associated with N3 target dropout (p<0.0001; RR: 31.03; 95% CI: 10.95-91.10). These results indicate that the G28916U and A28095U substitutions are associated with N2 and N3 target dropout, respectively, independent of quantity of SARS-CoV-2.

### Lineage-specific variation and target dropout

To determine whether target dropout was the result of lineage-specific variation, we examined phylogenetic lineages of genomes harboring the distinct polymorphisms. Among 25 specimens with N3 target dropout in this study, 23 (92%) belonged to the B.1.1.7 lineage (Alpha variant) and 2 were partial genomes whose phylogenies could not be determined. These specimens were collected from April 11, 2021 (PV36791) through June 30, 2021 (PV37690), which mirrored the prevalence of the Alpha variant in NYC (**Figure 1B**).

We next examined the lineage of genomes harboring the G28916U substitution (**Figure 4**). Of 111 specimens that bore the G28916U polymorphism and resulted in N2 target dropout, the earliest was from July 1, 2021 (PV37692) and the latest from August 21, 2021 (PV38720). All genomes corresponded with lineages and sublineages of the Delta variant. Specifically, these included the B.1.617.2 (n=67) lineage and AY.20 (n=2), AY.23 (n=1), AY.25 (n=22), AY.3 (n=17), AY.33 (n=1), and AY.5 (n=1) sublineages. We did not find the G28916U substitution among any of the other variant lineages in our dataset (**Figure 4A**).

**Figure 4.**
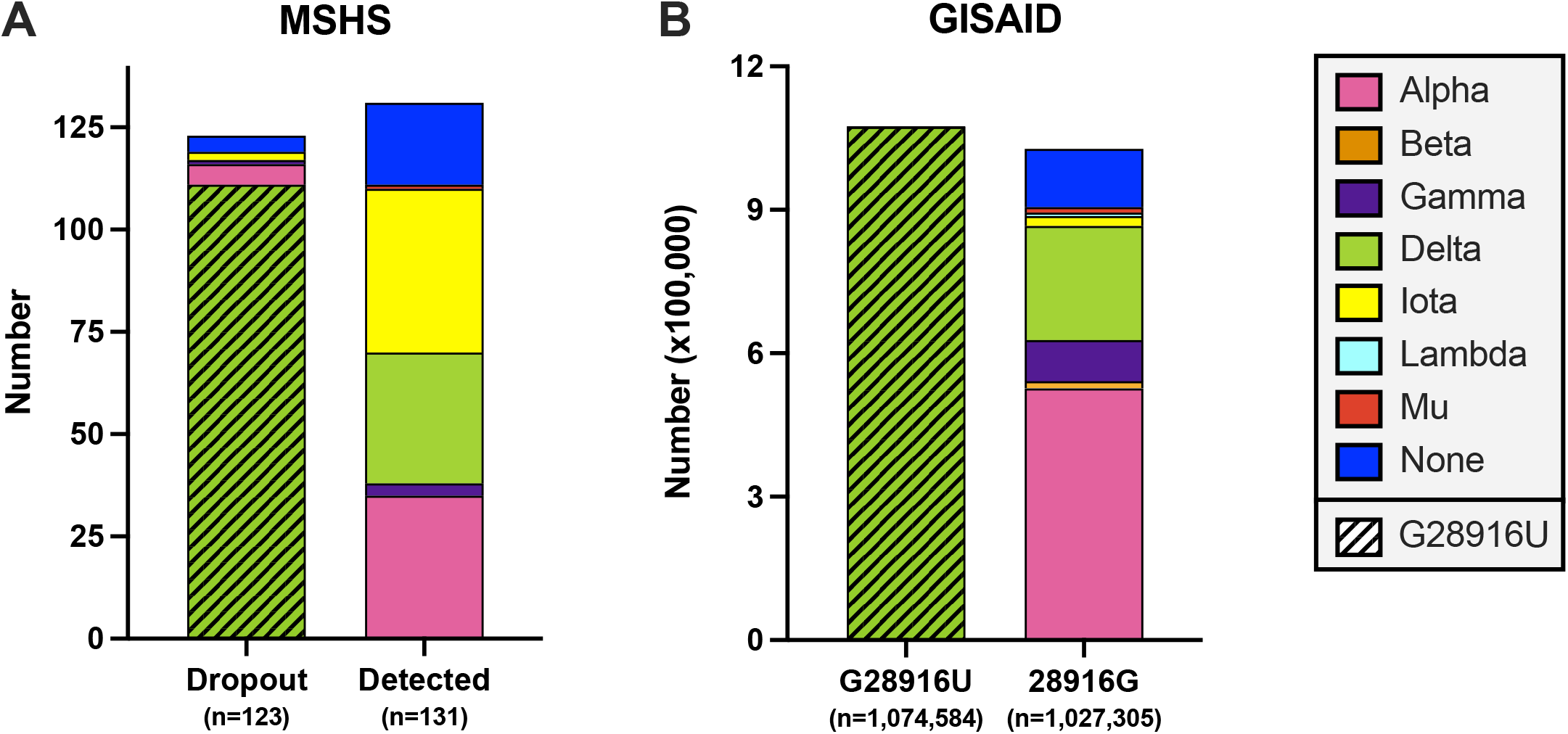
Delta-specific substitution interferes with N2 diagnostic target detection. (A) Stacked bar plots depict the composition of SARS-CoV-2 variants in 254 MSHS genomes tested by RT-PCR/MALDI-TOF. Bar plots reflect absolute number of genomes with N2 target dropout (left) and those with N2 target detection (right). Variant groups are color-coded and genomes with the G28916U polymorphism are depicted by a hatched pattern. (B) Stacked bar plot of publicly-available sequences (GISAID, see methods) from the same timeframe of this study depicting absolute numbers of variant genomes and presence or absence of the G28916U polymorphism. Color-coding and patterns are the same as in (A).

We broadened our analyses to interrogate SARS-CoV-2 genomes deposited globally for the G28916U polymorphism. We utilized the publicly-available EpiCoV (GISAID) database and queried 2,103,844 sequences collected during the same timeframe of this study (**Figure 4B**). We found the Delta variant (B.1.617.2 + AY.x) was predominant in the deposited genomes comprising 62% of all genomes (n=1,313,492). The majority of these Delta genomes (82%) harbored the G28916U substitution. Of the remaining Delta genome sequences, almost all (99.3%) had the native guanine nucleotide with a minority of sequences (<0.1%) harboring G28916A and G28916C substitutions. The G28916U polymorphism – which results in the G215C amino acid substitution in the N protein – is almost exclusive to Delta variant genomes but was rarely (<0.02%) reported in other variant groups including Alpha (B.1.1.7 + Q.x), Beta (B.1.351 + B.1.351.2 + B.1.351.3), Gamma (P.1 + P.1.x), Lambda (C.37 + C.37.1), and Mu (B.1.621 + B.1.621.1).

## Discussion

In the current study, we present a robust assessment of the impact of emergence of SARS-CoV-2 variants on detection of individual target genes included in the Agena MassARRAY^®^ SARS-CoV-2 Panel, focusing on a five-month period characterized by rapid changes in circulating VOC and VOI.

Molecular diagnostic tests to detect SARS-CoV-2, first designed almost two years ago, are challenged by continued viral circulation and rapid viral diversification, with continual emergence of new lineages. At the same time, recognition that some virus variants can demonstrate increased infectivity and disease severity among a broader population that now includes younger individuals ^2,13,16,17,44^ has raised awareness regarding assay sensitivity and risks for missed diagnoses. Although several studies have reported the impacts of viral sequence variation on conventional diagnostic platforms ^5,7,27,45^ or have highlighted, *in silico*, potential interference of variant polymorphisms with publicly-available target sequences ^42,46,47^, there are limited reports on assay performance *in actu* for detection of the Delta VOC and sublineages, and other recent VOC/VOI.

We utilized publicly-available primer/probe sequences of the Agena MassARRAY^®^ SARS-CoV-2 Panel and a deidentified dataset from clinical specimens that included individual target detection data and whole-genome sequencing to generate a snapshot of virus sequence diversity at each target PBS and to determine the extent to which polymorphisms are associated with individual target performance. Interestingly, we found that distinct viral substitutions resulted in N2 and N3 target dropout. Consistent with what we previously reported ^28^, the A28095U substitution is a B.1.1.7-associated polymorphism that introduces a mismatch at the 3’-end of the N3 forward PBS which results in N3 target dropout. Notably, a marked change in the pattern of target dropout occurred during the course of the study with the introduction of the Delta VOC; we found the G28916U polymorphism unique to a subset of Delta variants results in a 3’-mismatch to the N2 probe and is associated with N2 dropout. Our findings highlight the robust capability of this multiple-target platform to capture diverse viral variants, as well as the ability of target performance to reflect lineage-specific sequence variation outside the conventional *S* gene.

A number of recent reports have focused on variant panels designed to target a finite number of emergent mutations to discern variant identity, particularly in the first months of 2021. Most panels utilize RT-PCR or novel isothermal technologies (e.g., RT-LAMP, CRISPR/Cas-based, mass-spectrometry) to target specific polymorphisms in the *S* gene that have been ascribed to each of these variants ^25,27,45,48–50^. However, a recent deep analysis of circulating variants across the globe revealed that most of these signature mutations are not under positive selection ^1^. Therefore, other than a finite number of mutations in the spike protein (e.g., N501Y, S477N, V1176F, N501Y), one can anticipate that other variant-defining polymorphisms may not persist over continued circulation of SARS-CoV-2 and, thus, new target formulations will need to be continuously considered. As most efforts to accommodate new variants have been to redesign primers and probes or include more targets in this region ^45,51^, this may become impractical as inclusion of additional reactions can become costly and introduce more room for error. Furthermore, although multiplexing assays allow for single-pot reactions, the level of target redundancy is limited by the optical detection system as more fluorophores complicate conventional diagnostic methods ^52–54^. Therefore, as the COVID-19 pandemic continues and new variants emerge, there can be great utility in existing assay platforms that target multiple regions outside the *S* gene.

Our findings also highlight the level of sequence diversity observed at each of these five target loci among sequences from VOC/VOI detected through August 2021. Among the Agena MassARRAY^®^ SARS-CoV-2 Panel, *N* gene targets (e.g., N2, N3) are challenged by the greatest level of nucleotide diversity which can result in dropout of each. Consistent with phylogenetic analyses of global sequences ^1^, the N2 target region has the greatest sequence diversity. Although, a number of polymorphisms in the *N* gene are under positive selection ^1,46,55^, the evolutionary pressures exerted on A28095U (ORF8|K68*) and G28916U (N|G215C) polymorphisms have not demonstrated positive selection or purported epistatic interactions ^1,16^. This is likely due to the earlier timeframes for which each of these phylogenetic studies were performed (e.g., December 2020 through May 2021) and the lack of robust functional genomic studies in SARS-CoV-2. Thus, the fate of each of these polymorphisms is unclear among emergent VOC, and each should be taken into consideration for use of this diagnostic platform and others that target the same *N* gene regions going forward.

As SARS-CoV-2 variants continue to arise, robust platforms with high target redundancy for virus detection and diagnosis are vital to capture new or existing lineages. As contemporary polymorphisms undergo selection, evaluation of diagnostic target performance in the clinical setting can help elucidate ongoing viral dynamics in patient communities and highlight platform target combinations that should be further optimized. Lastly, our results reinforce the need for continuing surveillance of assay target result patterns to increase the sensitivity of current molecular tests for SARS-CoV-2 detection and to assure the adequate diagnosis of COVID-19 worldwide.

## Data Availability

To generate genome sequences, sequencing data were analyzed using a custom reference-based (MN908947.3) pipeline, https://github.com/mjsull/COVID_pipe. To analyze mismatches to diagnostic target PBSs, genome sequences were processed using a custom Unix-code https://github.com/AceM1188/SACOV_primer-probe_analyses.
SARS-CoV-2 sequencing read data for all study genomes were deposited in GISAID [www.gisaid.org] (accessions pending).

## Code availability

To generate genome sequences, sequencing data were analyzed using a custom reference-based (MN908947.3) pipeline, https://github.com/mjsull/COVID_pipe ^56^. To analyze mismatches to diagnostic target PBSs, genome sequences were processed using a custom Unix-code https://github.com/AceM1188/SACOV_primer-probe_analyses ^38^.

## Data availability

SARS-CoV-2 sequencing read data for all study genomes were deposited in GISAID [www.gisaid.org] (accessions pending).

## Acknowledgments

We thank the members of MSHS MML, Simon, and van Bakel laboratories for providing any assistance when needed throughout this study. We are grateful for the continuous expert guidance provided by the ISMMS Program for the Protection of Human Subjects (PPHS).

We are thankful for the efforts of the Mount Sinai Pathogen Surveillance Program (PSP) Study Group members: Hala Alshammary, Angela A. Amoako, Jose Polanco, Aria Rooker, Christian Cognigni, Mahmoud H. Awawda, Ashley-Beathrese T. Salimbangon, Zenab Khan, and Zain Khalil.

The Research reported in this paper was supported by the National Institutes of Health (NIH) contract number HHSN272201400008C, the NIH Office of Research Infrastructure under award numbers S10OD018522 and S10OD026880, institutional and philanthropic funds (Open Philanthropy Project, #2020-215611), as well as a Robin Chemers Neustein Postdoctoral Fellowship Award (to Dr. Gonzalez-Reiche).

## Author contributions

M.M.H., R.B., P.S., A.E.PM., M.R.G., M.D.N., and E.M.S. provided clinical samples for the study. M.M.H., R.B., B.G., P.S., L.C., F.C., H.S., A.H., M.S.PSP.S.G. and A.E.PM. accessioned clinical samples. A.S.GR., A.v.d.G, M.S.PSP.S.G., and H.v.B. performed NGS experiments. R.S. provided NGS services. A.S.GR. performed genome assembly, data curation and genotyping. M.M.H performed alignments and mismatch analyses. M.M.H., R.B., J.D.R., M.R.G., M.D.N., C.C.C., T.E.S., V.S., H.v.B., E.M.S., and A.E.PM analyzed, interpreted, or discussed data. M.M.H., E.M.S., and A.E.PM. wrote the manuscript. M.M.H., E.M.S., and A.E.PM. conceived the study. E.M.S. and A.E.PM. supervised the study. H.v.B., V.S., and E.M.S. raised financial support.

M.M.H. and A.E.PM are the guarantors of this work and, as such, had full access to all of the data in the study and take responsibility for the integrity of the data and the accuracy of the data analysis.

