## Supplemental Info for "RT-PCR/MALDI-TOF diagnostic target performance reflects circulating SARS-CoV-2 variant diversity in New York City"

**SUPPLEMENTAL DATA**

**Table S1. Agena MassARRAY^®^ RT-PCR/MALDI-TOF target primer and probe sequences**

| **Target** | **Primer/Probe** | **Sequence (5’->3’)** | **SARS-CoV-2 Coordinates (NC_045512.2)** |
| --- | --- | --- | --- |
| N1 | For | AGACGGCATCATATGGGTTG | 28654 - 28673 |
| N1 | Rev | TGAGGAAGTTGTAGCACGATTG | 28761 - 28740 |
| N1 | Probe | GTGCCAATGTGATCTTT | 28716 - 28700 |
| N2 | For | GGGGAACTTCTCCTGCTAGAAT | 28881 - 28902 |
| N2 | Rev | CAGACATTTTGCTCTCAAGCTG | 28979 - 28958 |
| N2 | Probe | GCAAAGCAAGAGCAGCATCACC | 28937 - 28916 |
| N3 | For | GTGGATGAGGCTGGTTCTAA | 28096 - 28077 |
| N3 | Rev | ACTACAAGACTACCCAATTT | 28192 - 28173 |
| N3 | Probe | GAAACTGTATAATTACCGATA | 28138 - 28118 |
| ORF1A | For | AACTGTTGGTCAACAAGACG | 3223 - 3242 |
| ORF1A | Rev | CAATAGTCTGAACAACTGGTGT | 3335 - 3314 |
| ORF1A | Probe | GGTTCAACCTCAATTAG | 3286 - 3302 |
| ORF1AB | For | CCCTGTGGGTTTTACACTTAA | 13342 - 13362 |
| ORF1AB | Rev | ACGATTGTGCATCAGCTGA | 13460 - 13442 |
| ORF1AB | Probe | ATCAACTCCGCGAACCC | 13432 - 13416 |

**Table S2. Targets detected for individual specimens.**

| **Specimen ID** | **N1*** | **N2*** | **N3*** | **ORF1A*** | **ORF1AB*** |
| --- | --- | --- | --- | --- | --- |
| PV36456 | D | ND | D | D | D |
| PV36458 | D | D | D | D | ND |
| PV36791 | D | ND | ND | D | ND |
| PV36644 | D | D | D | D | D |
| PV36640 | D | D | D | D | D |
| PV36645 | D | D | D | D | D |
| PV36639 | D | D | D | D | D |
| PV36648 | D | D | ND | D | D |
| PV36646 | D | D | D | D | D |
| PV36641 | D | D | D | D | D |
| PV36642 | D | D | D | D | D |
| PV36647 | D | D | D | D | D |
| PV36649 | D | D | D | D | D |
| PV36652 | D | ND | ND | D | ND |
| PV36653 | D | D | D | D | D |
| PV36654 | ND | D | D | D | D |
| PV36650 | D | D | D | D | ND |
| PV36651 | D | D | D | D | D |
| PV36655 | D | D | D | D | D |
| PV36656 | D | D | D | D | D |
| PV36659 | D | D | D | D | D |
| PV36660 | D | D | D | D | D |
| PV36661 | D | D | D | D | D |
| PV36663 | D | ND | D | D | D |
| PV36664 | D | D | ND | D | D |
| PV36666 | D | D | D | D | D |
| PV36670 | D | D | D | D | ND |
| PV36671 | D | D | D | D | D |
| PV36672 | D | D | D | D | D |
| PV36673 | D | D | D | D | D |
| PV36674 | D | D | D | D | D |
| PV36677 | D | D | D | D | D |
| PV36678 | D | D | ND | D | D |
| PV36689 | D | D | D | D | D |
| PV36681 | D | D | ND | D | D |
| PV36682 | D | D | D | D | D |
| PV36685 | D | D | ND | D | D |
| PV36684 | D | D | D | D | D |
| PV36683 | D | D | ND | D | ND |
| PV36690 | D | D | D | D | D |
| PV36692 | D | D | D | D | D |
| PV36889 | D | D | D | D | D |
| PV36890 | D | D | ND | D | D |
| PV36924 | D | D | D | D | D |
| PV36925 | D | D | D | D | D |
| PV36931 | D | D | D | D | D |
| PV36932 | D | D | D | D | D |
| PV36940 | D | D | D | D | D |
| PV36951 | D | D | ND | D | D |
| PV36945 | D | D | D | D | D |
| PV36946 | D | D | ND | D | D |
| PV36947 | D | D | D | D | D |
| PV36948 | D | D | D | D | D |
| PV36949 | D | D | D | D | D |
| PV36950 | D | D | D | D | ND |
| PV37050 | D | D | ND | D | D |
| PV37051 | D | D | D | D | D |
| PV37052 | D | D | ND | D | D |
| PV37074 | D | D | ND | D | D |
| PV37054 | D | D | D | D | ND |
| PV37057 | D | D | ND | D | D |
| PV37058 | D | D | ND | D | ND |
| PV37060 | D | D | D | D | D |
| PV37059 | D | D | ND | ND | ND |
| PV37061 | D | ND | ND | D | ND |
| PV37066 | D | D | D | D | D |
| PV37067 | D | ND | ND | D | D |
| PV37062 | D | D | ND | D | D |
| PV37064 | D | D | ND | D | ND |
| PV37065 | D | D | D | D | D |
| PV37069 | D | D | D | D | D |
| PV37070 | D | ND | D | D | ND |
| PV37073 | D | D | D | D | D |
| PV37267 | D | D | D | D | D |
| PV37269 | D | D | D | D | D |
| PV37270 | D | D | D | D | D |
| PV37272 | D | D | ND | D | ND |
| PV37271 | D | D | ND | D | D |
| PV37273 | D | D | ND | D | D |
| PV37274 | D | D | D | D | D |
| PV37276 | D | D | ND | D | ND |
| PV37277 | D | D | D | ND | ND |
| PV37275 | D | D | D | D | D |
| PV37282 | D | D | D | D | D |
| PV37283 | D | D | ND | D | D |
| PV37284 | D | ND | ND | D | ND |
| PV37281 | D | D | D | D | D |
| PV37286 | D | D | D | D | D |
| PV37288 | D | D | D | D | D |
| PV37292 | D | D | D | D | D |
| PV37295 | D | D | D | D | D |
| PV37290 | D | D | ND | D | D |
| PV37291 | D | D | ND | D | D |
| PV37296 | D | ND | D | D | D |
| PV37297 | D | D | D | D | ND |
| PV37300 | D | D | D | D | D |
| PV37303 | D | ND | D | D | ND |
| PV37301 | D | D | D | D | D |
| PV37293 | D | D | ND | D | D |
| PV37463 | D | D | D | D | D |
| PV37467 | D | D | D | D | D |
| PV37468 | D | D | D | D | D |
| PV37471 | D | D | D | D | D |
| PV37501 | D | ND | D | D | ND |
| PV37499 | D | ND | D | D | D |
| PV37509 | D | D | ND | D | D |
| PV37633 | D | D | D | D | D |
| PV37652 | D | D | D | D | D |
| PV37650 | D | D | D | D | D |
| PV37651 | D | D | D | D | D |
| PV37666 | D | D | D | D | D |
| PV37690 | D | D | ND | D | D |
| PV37692 | D | ND | D | D | D |
| PV37689 | D | D | D | D | D |
| PV37986 | D | ND | D | D | D |
| PV37985 | D | D | D | D | D |
| PV38040 | D | ND | ND | D | ND |
| PV38041 | D | D | D | D | D |
| PV38161 | D | D | D | D | D |
| PV38160 | D | ND | D | D | D |
| PV33505 | D | ND | D | D | D |
| PV33516 | D | ND | D | D | D |
| PV33515 | D | ND | D | D | D |
| PV33517 | D | ND | D | D | D |
| PV33518 | D | ND | D | D | D |
| PV33511 | D | ND | D | D | D |
| PV33503 | D | ND | D | D | D |
| PV33504 | D | D | D | D | D |
| PV33502 | D | ND | D | D | D |
| PV33513 | D | ND | D | D | D |
| PV33522 | D | ND | D | D | D |
| PV33521 | D | ND | D | D | D |
| PV33523 | D | ND | D | D | D |
| PV38267 | D | ND | D | D | D |
| PV38268 | D | ND | D | D | D |
| PV38270 | D | ND | D | D | D |
| PV38272 | D | D | D | D | D |
| PV38273 | D | D | D | D | D |
| PV38288 | D | ND | D | D | D |
| PV38269 | D | ND | D | D | D |
| PV38347 | D | ND | D | D | D |
| PV38289 | D | ND | D | D | D |
| PV38276 | D | D | D | D | D |
| PV38277 | D | D | D | D | D |
| PV38279 | D | ND | D | D | D |
| PV38278 | D | ND | D | D | D |
| PV38282 | D | ND | D | D | D |
| PV38283 | D | ND | D | D | D |
| PV38284 | D | ND | D | D | D |
| PV38285 | D | ND | D | D | D |
| PV38286 | D | D | D | D | D |
| PV38287 | D | D | D | D | D |
| PV38346 | D | ND | D | D | D |
| PV38407 | D | ND | D | D | D |
| PV38356 | D | ND | D | D | D |
| PV38348 | D | ND | D | D | D |
| PV38343 | D | ND | D | D | D |
| PV38344 | D | D | D | D | D |
| PV38345 | D | ND | D | D | D |
| PV38335 | D | D | D | D | D |
| PV38337 | D | ND | D | D | D |
| PV38338 | D | ND | D | D | D |
| PV38340 | D | ND | D | D | D |
| PV38351 | D | D | D | D | D |
| PV38350 | D | ND | D | D | D |
| PV38352 | D | D | D | D | D |
| PV38359 | D | ND | D | D | D |
| PV38405 | D | ND | D | D | D |
| PV38360 | D | D | D | D | D |
| PV38361 | D | D | D | D | D |
| PV38363 | D | ND | D | D | D |
| PV38353 | D | ND | D | D | D |
| PV38354 | D | ND | D | D | D |
| PV38355 | D | ND | D | D | D |
| PV38365 | D | D | D | D | D |
| PV38366 | D | ND | D | D | D |
| PV38368 | D | ND | D | D | D |
| PV38372 | D | D | D | D | D |
| PV38369 | D | D | D | D | D |
| PV38373 | D | ND | D | D | D |
| PV38370 | D | ND | D | D | D |
| PV38374 | D | D | D | D | D |
| PV38371 | D | D | D | D | D |
| PV38376 | D | ND | D | D | ND |
| PV38377 | D | ND | D | D | D |
| PV38378 | D | ND | D | D | D |
| PV38384 | D | D | D | D | D |
| PV38383 | D | ND | D | D | D |
| PV38385 | D | ND | D | D | D |
| PV38386 | D | ND | D | D | D |
| PV38387 | D | ND | D | D | D |
| PV38393 | D | ND | D | D | D |
| PV38391 | D | ND | D | D | D |
| PV38394 | D | ND | D | D | D |
| PV38392 | D | ND | D | D | D |
| PV38389 | D | D | D | D | D |
| PV38399 | D | ND | D | D | D |
| PV38400 | D | ND | D | D | D |
| PV38401 | D | ND | D | D | D |
| PV38402 | D | D | D | D | D |
| PV38403 | D | ND | D | D | D |
| PV38404 | D | ND | D | D | D |
| PV38390 | D | ND | D | D | D |
| PV38395 | D | ND | D | D | D |
| PV38398 | D | ND | D | D | D |
| PV38681 | D | D | D | D | D |
| PV38686 | D | D | D | D | D |
| PV38680 | D | D | D | D | D |
| PV38672 | D | ND | D | D | D |
| PV38688 | D | ND | D | D | D |
| PV38659 | D | ND | D | D | D |
| PV38660 | D | D | D | D | D |
| PV38666 | D | ND | D | D | D |
| PV38693 | D | ND | D | D | D |
| PV38685 | D | ND | D | D | D |
| PV38664 | D | ND | D | D | D |
| PV38676 | D | ND | D | D | ND |
| PV38656 | D | ND | D | D | D |
| PV38674 | D | ND | D | D | D |
| PV38655 | D | ND | D | D | D |
| PV38694 | D | ND | D | D | D |
| PV38663 | D | ND | D | D | D |
| PV38682 | D | ND | D | D | D |
| PV38662 | D | ND | D | D | D |
| PV38683 | D | D | D | D | D |
| PV38691 | D | D | D | D | D |
| PV38695 | D | ND | D | D | D |
| PV38675 | D | ND | D | D | D |
| PV38689 | D | ND | D | D | D |
| PV38692 | D | ND | D | D | D |
| PV38677 | D | ND | D | D | D |
| PV38684 | D | ND | D | D | D |
| PV38657 | D | ND | D | D | D |
| PV38667 | D | ND | D | D | D |
| PV38661 | D | ND | D | D | D |
| PV38750 | D | ND | D | D | D |
| PV38678 | D | ND | D | D | D |
| PV38670 | D | ND | D | D | D |
| PV38665 | D | ND | ND | D | ND |
| PV38699 | D | ND | D | D | D |
| PV38715 | D | ND | ND | D | ND |
| PV38707 | D | ND | D | D | D |
| PV38698 | D | ND | D | D | D |
| PV38713 | D | ND | D | D | D |
| PV38706 | D | ND | D | D | D |
| PV38701 | D | ND | D | D | D |
| PV38668 | D | ND | D | D | D |
| PV38718 | D | ND | D | D | D |
| PV38712 | D | ND | D | D | D |
| PV38705 | D | ND | D | D | D |
| PV38703 | D | ND | D | D | D |
| PV38722 | D | ND | D | D | D |
| PV38697 | D | ND | D | D | D |
| PV38716 | D | ND | D | D | D |
| PV38720 | D | ND | D | D | D |
| PV38840 | D | D | ND | D | D |
| *Detected, D; Not detected, ND | | | | | |

**Figure S1. Positions of PBS mismatches and target detection results.** Line graphs depict the percentage of specimen genomes with mismatches at individual positions across the (A) N1, (B) N2, (C) N3, (D) ORF1A, and (E) ORF1AB target PBSs. Forward (For), Reverse (Rev), and Probe PBS are depicted in separate plots for each target. For each PBS, two line plots depict mismatches in genomes from specimens that yielded a detected target result (magenta) or target dropout (turquoise). The percentage represents the number of genomes with mismatches at each position relative to the number of genome sequences detected or not detected by each target (annotated in each graph).
