## Supplementary figures and images for "RT-PCR/MALDI-TOF diagnostic target performance reflects circulating SARS-CoV-2 variant diversity in New York City"

### Supplemental Figures

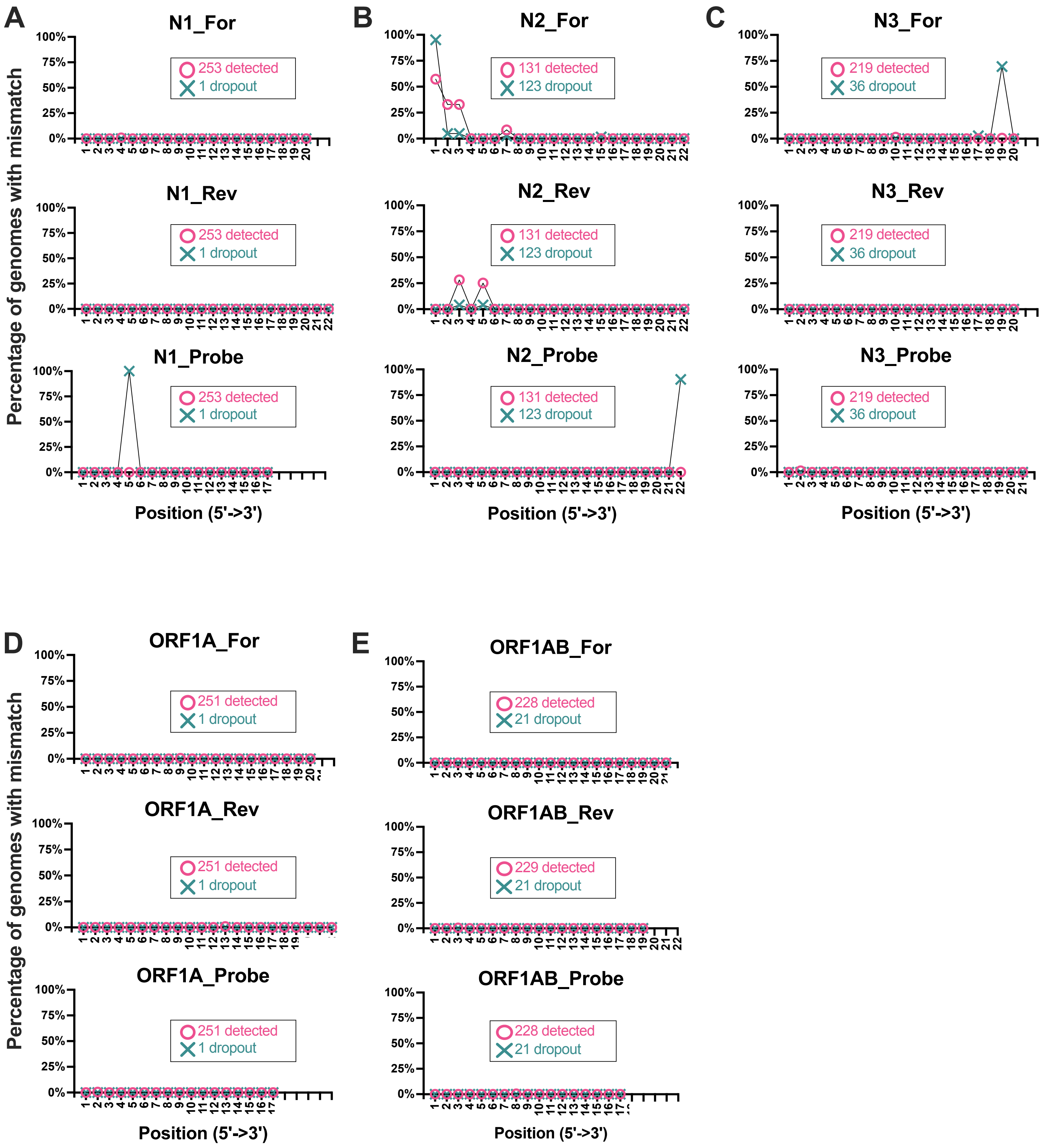
